# Relationship between the ABO Blood Group and the COVID-19 Susceptibility

**DOI:** 10.1101/2020.03.11.20031096

**Authors:** Jiao Zhao, Yan Yang, Hanping Huang, Dong Li, Dongfeng Gu, Xiangfeng Lu, Zheng Zhang, Lei Liu, Ting Liu, Yukun Liu, Yunjiao He, Bin Sun, Meilan Wei, Guangyu Yang, Xinghuan Wang, Li Zhang, Xiaoyang Zhou, Mingzhao Xing, Peng George Wang

## Abstract

The novel coronavirus disease-2019 (COVID-19) has been spreading around the world rapidly and declared as a pandemic by WHO. Here, we compared the ABO blood group distribution in 2,173 patients with COVID-19 confirmed by SARS-CoV-2 test from three hospitals in Wuhan and Shenzhen, China with that in normal people from the corresponding regions. The results showed that blood group A was associated with a higher risk for acquiring COVID-19 compared with non-A blood groups, whereas blood group O was associated with a lower risk for the infection compared with non-O blood groups. This is the first observation of an association between the ABO blood type and COVID-19. It should be emphasized, however, that this is an early study with limitations. It would be premature to use this study to guide clinical practice at this time, but it should encourage further investigation of the relationship between the ABO blood group and the COVID-19 susceptibility.

## INTRODUCTION

The novel coronavirus SARS-CoV-2, causing the new infectious coronavirus disease-2019 (COVID-19), is currently spreading rapidly around the world; it has been recently declared as a pandemic by WHO. Recent clinical observation suggests that patient age, male sex and certain chronic medical conditions (e.g., cardiovascular disease, diabetes, COPD) seem to represent a risk for the infection of SARS-Cov-2 and higher disease severity^1^. There is currently no biological marker known to predict the susceptibility to COVID-19. Landsteiner’s ABO blood types are carbohydrate epitopes that are present on the surface of human cells. The antigenic determinants of A and B blood groups are trisaccharide moieties GalNAcα1-3-(Fucα1,2)-Galβ- and Galα1-3-(Fucα1,2)-Galβ-, while O blood group antigen is Fucα1,2-Galβ-. While blood types are genetically inherited, the environment factors can potentially influence which blood types in a population will be passed on more frequently to the next generation. Susceptibility of viral infection has been previously found to be related to ABO blood group. For example, Norwalk virus and Hepatitis B have clear blood group susceptibility^2,3^. It was also reported that blood group O individuals were less likely to become infected by SARS coronavirus^4^. Here, we investigated the relationship between the ABO blood type and the susceptibility to COVID-19 in patients from three hospitals in Wuhan and Shenzhen, China to test if the former may be a biomarker for the latter.

## METHODS

We collected and ABO-typed blood samples from 1,775 patients infected with SARS-CoV-2, including 206 dead cases, at the Jinyintan Hospital in Wuhan, Hubei province, China. Another 113 and 285 patients with COVID-19 were respectively recruited from Renmin Hospital of Wuhan University, Hubei province and Shenzhen Third People’s Hospital, Guangdong province, China. The diagnosis of COVID-19 was confirmed by a positive real-time reverse transcriptase polymerase-chain-reaction test of SARS-CoV-2 on nasal and pharyngeal swab specimens from patients. Two recent surveys of ABO blood group distribution of 3,694 normal people from Wuhan City and 23,386 normal people from Shenzhen City were used as comparison controls for the Wuhan and Shenzhen patients with COVID-19, respectively^5-6^. Statistical analyses were performed using chi-squared test. Data from different hospitals were meta-analyzed using random effects models, with calculation of odds ratio (OR) and 95% confidence interval (*CI*). Statistical analyses were performed using SPSS software (version 16.0) and STATA software (version 13).

## RESULTS

The ABO blood group in 3,694 normal people in Wuhan displayed a percentage distribution of 32.16%, 24.90%, 9.10% and 33.84% for A, B, AB and O, respectively, while the 1,775 patients with COVID-19 from Wuhan Jinyintan Hospital showed an ABO distribution of 37.75%, 26.42%, 10.03% and 25.80% for A, B, AB and O, respectively. The proportion of blood group A in patients with COVID-19 was significantly higher than that in normal people, being 37.75% in the former vs 32.16% in the later (*P* < 0.001). The proportion of blood group O in patients with COVID-19 was significantly lower than that in normal people, being 25.80% in the former vs 33.84% in the later (P < 0.001, Table 1). These results corresponded to a significantly increased risk of blood group A for COVID-19 with an OR of 1.279 (95% *CI* 1.136∼1.440) and decreased risk of blood group O for COVID-19 with an OR of 0.680 (95% *CI* 0.599∼0.771, Table 1) in comparison with non-A groups and non-O groups, respectively.

**Table 1.** The ABO blood group distribution in patients with COVID-19 and normal controls.

|  | Blood Group |  |  |  |
| --- | --- | --- | --- | --- |
|  | A | B | AB | O |
| Controls (Wuhan Area) |  |  |  |  |
| 3694 | 1188 (32.16%) | 920 (24.90%) | 336 (9.10%) | 1250 (33.84%) |
| **Wuhan Jinyintan Hospital** |  |  |  |  |
| Patients |  |  |  |  |
| 1775 | 670 (37.75%) | 469 (26.42%) | 178 (10.03%) | 458 (25.80%) |
| $\chi^2$ | 16.431 | 1.378 | 1.117 | 35.674 |
| *P* | **<0.001** | 0.240 | 0.291 | **<0.001** |
| OR | 1.279 | 1.083 | 1.114 | 0.680 |
| 95%CI | 1.136~1.440 | 0.952~1.232 | 0.920~1.349 | 0.599~0.771 |
| Deaths |  |  |  |  |
| 206 | 85 (41.26%) | 50 (24.27%) | 19 (9.22%) | 52 (25.24%) |
| $\chi^2$ | 6.944 | 0.015 | 0.000 | 6.102 |
| *P* | **0.008** | 0.903 | 1.000 | **0.014** |
| OR | 1.482 | 0.966 | 1.015 | 0.660 |
| 95%CI | 1.113~1.972 | 0.697~1.340 | 0.625~1.649 | 0.479~0.911 |
| **Renmin Hospital of Wuhan University** |  |  |  |  |
| 113 | 45 (39.82%) | 25 (22.12%) | 15 (13.3%) | 28 (24.78%) |
| patients |  |  |  |  |
| $\chi^2$ | 2.601 | 0.318 | 1.815 | 3.640 |
| *P* | 0.107 | 0.573 | 0.178 | 0.045 |
| OR | 1.396 | 0.857 | 1.530 | 0.644 |
| 95%CI | 0.952~2.048 | 0.546~1.344 | 0.878~2.664 | 0.418~0.993 |
| Controls (Shenzhen area) |  |  |  |  |
| 23386 | 6728 (28.77%) | 5880 (25.14%) | 1712 (7.32%) | 9066 (38.77%) |
| **Patients from Shenzhen Third People's Hospital** |  |  |  |  |
| 285 | 82 (28.77%) | 83 (29.12%) | 39 (13.68%) | 81 (28.42%) |
| $\chi^2$ | 0.000 | 2.160 | 15.729 | 12.278 |
| *P* | 1.000 | 0.142 | **<0.001** | 0.001 |
| OR | 1.000 | 1.223 | 2.008 | 0.627 |
| 95%CI | 0.773~1.294 | 0.946~1.582 | 1.427~2.824 | 0.484~0.812 |

CI, confidence interval; OR, odds ratio; \**P* value was calculated by 2-tailed $\chi^2$

A similar distribution pattern of high-risk blood group A and low-risk blood group O was also observed in the dead patients. Specifically, the proportions of blood groups A, B, AB and O in the 206 dead patients were 41.26%, 24.27%, 9.22% and 25.24%, respectively. Blood group A was associated with a higher risk of death compared with non-A groups, with an OR of 1.482 (95% *CI* 1.113∼1.972, *P* = 0.008, Table 1). To the contrary, blood group O was associated with a lower risk of death compared with non-O groups, with an OR of 0.660 (95% *CI* 0.479∼0.911, *P* = 0.014, Table 1).

We next examined 113 patients with COVID-19 from another hospital in Wuhan City, the Renmin Hospital of Wuhan University, and found a similar risk distribution trend of ABO blood groups for the infection. Specifically, compared with non-A blood groups, blood group A displayed higher relative risk (OR=1.396; 95% *CI* 0.952∼2.048) than those observed in patients from Wuhan Jinyintan Hospital, although the associations did not reach statistical significance likely due to the small sample size. Compared with non-O groups, blood group O were significantly associated with a lower risk of infection, with an OR of 0.644 (95% *CI* 0.418∼0.993, *P* = 0.045, Table 1).

The ABO blood group in 23,368 normal people in Shenzhen displayed a percentage distribution of 28.77%, 25.14%, 7.32% and 38.77% for A, B, AB and O, respectively. Analysis of 285 patients with COVID-19 from Shenzhen showed proportions of blood groups A, B, AB and O to be 28.77%, 29.12%, 13.68% and 28.42%, respectively. These results similarly showed a significantly lower risk of infection associated with blood group O (OR, 0.627; 95% *CI* 0.484∼0.812). These results also showed blood group AB to have an increased risk of infection (OR, 2.008; 95% *CI* 1.427∼2.824, Table 1).

Figure 1 shows the estimates of ORs of the risk of ABO blood groups for COVID-19 on the pooled data from the three hospitals by random effects models. Again, the results showed that blood group A was associated with a significantly higher risk for COVID-19 (OR, 1.21; 95% *CI* 1.02∼1.43, *P* = 0.027) compared with non-A blood groups, whereas blood group O was associated with a significantly lower risk for the infection (OR, 0.67; 95% *CI* 0.60∼0.75, *P* < 0.001) compared with non-O blood groups. Compared with other ABO blood groups, AB blood group (OR, 1.48, 95% *CI* 0.97∼2.24) and B blood group (OR, 1.09, 95% *CI* 0.98∼1.22) seemed to have a trend of higher risk for infection, but the association did not reach statistical significance.

**Figure 1.**
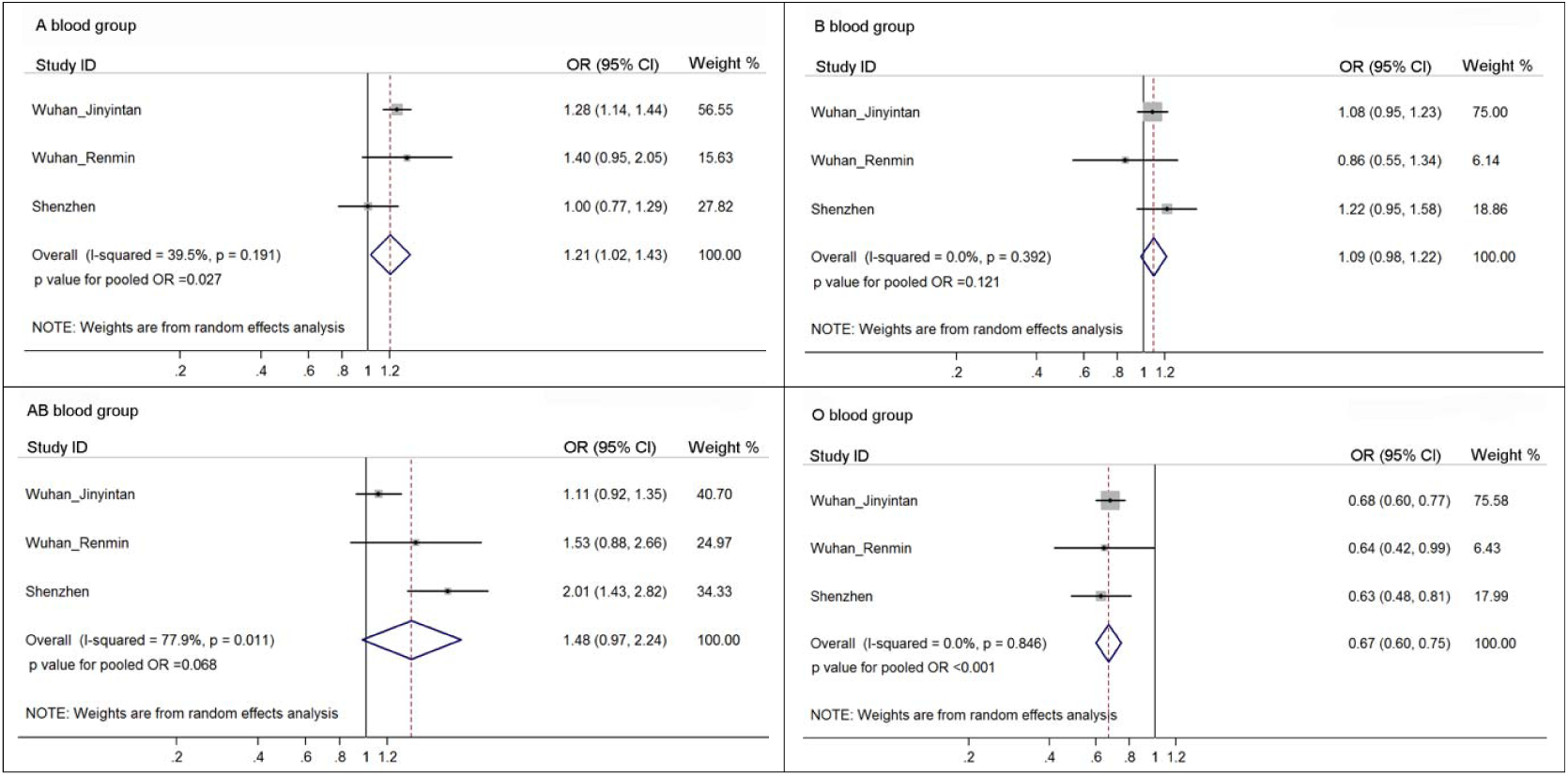
Meta-analysis of the risk of ABO blood groups for COVID-19 in three hospitals. The X-axis represents the point estimate of odds ratio and corresponding 95% confidence interval; the Y-axis represents the source of study patients. OR, odds ratio. *CI*, confidence interval.

We next investigated whether patient age and sex might influence the ABO blood group distribution among patients with COVID-19. We used the ABO blood group distribution of 3,694 normal people in Wuhan area as control to compare with different age and sex groups. When all patients from Jinyintan Hospital and Renmin Hospital in Wuhan city were combined (1,888 patients in total) and grouped into three age groups (≤40, 41-59, ≥60 years old), the ABO blood group distribution was similar among the three age groups (Table S1). The ABO blood group distribution was also similar between male and female patients with COVID-19 (Table S1). These results are consistent with the fact that the distribution of ABO blood groups is known to have no sex and age predilections. For example, by analyzing the blood type of more than ninety thousands normal people, it was found that the percentage of A, B, AB and O blood types were essentially the same among different age groups and among different sexes.^7^

## DISCUSSION

In this study, we found that ABO blood groups displayed different association risks for the infection with SARS-CoV-2 resulting in COVID-19. Specifically, blood group A was associated with an increased risk whereas blood group O was associated with a decreased risk, thus demonstrating that the ABO blood type is a biomarker for differential susceptibility of COVID-19. These findings are consistent with similar risk patterns of ABO blood groups for other coronavirus infection found in previous studies. For example, Cheng *et al*. reported that the SARS-CoV infection susceptibility in Hong Kong was differentiated by the ABO blood group systems^4^. The authors found that compared with non-O blood group hospital staff, blood group O hospital staff had a lower chance of getting infected. Patrice *et al*. found that anti-A antibodies specifically inhibited the adhesion of SARS-CoV S protein-expressing cells to ACE2-expressing cell lines^7^. Given the nucleic acid sequence similarity^8^ and receptor angiotensin-converting enzyme 2 (ACE2) binding similarity between SARS-CoV and SARS-CoV-2^9-11^, the lower susceptibility of blood group O and higher susceptibility of blood group A for COVID-19 could be linked to the presence of natural anti-blood group antibodies, particularly anti-A antibody, in the blood. This speculation will need direct studies to prove. There may also be other mechanisms underlying the ABO blood group-differentiated susceptibility for COVID-19 that require further studies to elucidate.

### Limitations

This study has several limitations. 1) The numbers of patients from the Renmin Hospital of Wuhan University and Shenzhen Third People’s Hospital were both small. Therefore, they cannot represent a solid replication analysis, although the data from these two hospitals showed similar risk patterns of ABO blood groups. 2) The control population groups used in this study lacked information on the subject age and sex and, therefore, a multivariate analysis to adjust the effect of the two factors was not possible. This, however, may not necessarily be a major issue since the ABO blood group distribution was similar among different age and sex groups of patients in the present study and in normal population reported previously^7^. 3) Due to incomplete information, the influence of the status of chronic medical conditions, such as vascular disease, diabetes mellitus and COPD, could not be adjusted through a multivariate analysis, which could potentially bias the conclusions of the present study since these factors may affect the severity of COVID-19.

### Conclusions

In this novel study, we for the first time report an association between the ABO blood group and COVID-19 susceptibility, demonstrating the latter to be a biomarker differentiating the former. Specifically, people with blood group A have a higher risk whereas people with blood group O have a lower risk for SARS-Cov-2 infection and COVID-19 severity. If verified by future studies, the findings in the present study would have several potential clinical implications. 1) People with blood group A might need particularly strengthened personal protection to reduce the chance of infection; 2) SARS-CoV-2-infected patients with blood group A might need to receive more vigilant surveillance and aggressive treatment; 3) It might be helpful to introduce ABO blood typing in the management of SARS-CoV-2 infection and COVID-19.

It should be emphasized, however, that given the above limitations, it would be premature to use this study to guide clinical practice at this time. Large replication studies with complete information should be encouraged to pursue and are needed to verify the present findings. Obviously, people with any blood type all need to exercise the wisdom of careful practice to avoid SARS-CoV-2 infection.

## Data Availability

The data used to support the findings of this study are included within the article.

## Author contributions statement

P.G.W and G.Y.Y conceived, designed and supervised the overall study. P.G.W, M.X, G.Y.Y, L.Z, and X.Y.Z supervised and administered the project. L.Z, H.P.H and T.L collected and verified ABO blood types of patients from Wuhan Jinyintan Hospital. X.Y.Z and D.L collected and verified ABO blood types of patients from Renmin Hospital of Wuhan University. Z.Z, L.L and Y.Y collected and verified ABO blood types of patients from Second Affiliated Hospital, Southern University of Science and Technology. Y.J.H, B.S, M.L.W, X.H.W collected and verified the data. D.F.G, X.F.L, Y.K.L, Z.J, M.X, and P.G.W analyzed the data. P.G.W, J.Z, G.Y.Y and M.X, wrote and revised the paper. All authors read and approved the final manuscript.

## PATIENTS AND PUBLIC INVOLVEMENT

This was a retrospective case series study and no patients were directly involved in the study design, setting the research questions, or the outcome measures directly. No patients were asked to advise on interpretation or writing up of results.

## Funding

No funding.

## Competing interest statement

The authors declare that they have no competing financial interests.

## Ethical approval

This study received approval from the Research Ethics Committees of the participating institutions, which waivered informed patient consent because the study only involved retrospective review of clinical data and because of the urgent nature of the study to investigate a new serious infections disease.

## Patient consent

Waived

## Data sharing

No additional data available.

## Data Availability Statement

The data used to support the findings of this study are included within the article.

---

**Table S1.** Influence of age and gender on the ABO blood group distribution in patients with COVID-19 combined from two Wuhan hospitals.

|  | Blood Group |  |  |  |
| --- | --- | --- | --- | --- |
|  | A | B | AB | O |
| 3694 Control (Wuhan area) | 1188 (32.16%) | 920 (24.90%) | 336 (9.10%) | 1250 (33.84%) |
| **Wuhan** |  |  |  |  |
| 1888 patients | 715 (37.87%) | 494 (26.17%) | 193 (10.22%) | 486 (25.74%) |
| $\chi^2$ | 17.880 | 0.983 | 1.720 | 37.852 |
| *P* | **<0.001** | 0.321 | 0.190 | **<0.001** |
| OR | 1.286 | 1.069 | 1.138 | 0.678 |
| 95%CI | 1.145~1.444 | 0.941~1.213 | 0.944~1.371 | 0.599~0.767 |
| Less than 40 years | 100 (36.63%) | 78 (28.57%) | 21 (7.69%) | 74 (27.11%) |
| $\chi^2$ | 2.117 | 1.625 | 0.452 | 4.883 |
| *P* | **0.146** | 0.202 | 0.501 | **0.027** |
| OR | 1.219 | 1.206 | 0.833 | 0.727 |
| 95%CI | 0.944~1.575 | 0.918~1.585 | 0.526~1.318 | 0.552~0.958 |
| Between 41-59 years | 275 (39.01%) | 176 (24.96%) | 71 (10.07%) | 183 (25.96%) |
| $\chi^2$ | 12.197 | <0.001 | 0.559 | 16.385 |
| *P* | **<0.001** | 1.000 | 0.455 | **<0.001** |
| OR | 1.349 | 1.003 | 1.119 | 0.685 |
| 95%CI | 1.142~1.593 | 0.833~1.208 | 0.855~1.466 | 0.572~0.822 |
| Over 60 years | 340 (37.36%) | 240 (26.37%) | 101 (11.10%) | 229 (25.16%) |
| $\chi^2$ | 8.679 | 0.759 | 3.181 | 24.797 |
| *P* | **0.003** | 0.384 | **0.075** | **<0.001** |
| OR | 1.258 | 1.080 | 1.248 | 0.657 |
| 95%CI | 1.082~1.463 | 0.916~1.274 | 0.986~1.579 | 0.558~0.775 |
| male 1030 | 403 (39.13%) | 275 (26.70%) | 101 (9.81%) | 251 (24.37%) |
| $\chi^2$ | 17.187 | 1.278 | 0.403 | 32.883 |
| *P* | **<0.001** | 0.258 | 0.526 | **<0.001** |
| OR | 1.356 | 1.098 | 1.087 | 0.630 |
| 95%CI | 1.175~1.564 | 0.939~1.285 | 0.860~1.373 | 0.538~0.738 |
| female 858 | 312 (36.36%) | 219 (25.52%) | 92 (10.72%) | 235 (27.39%) |
| $\chi^2$ | 5.379 | 0.111 | 1.976 | 12.884 |
| *P* | **0.020** | 0.739 | 0.160 | **<0.001** |
| OR | 1.205 | 1.033 | 1.200 | 0.738 |
| 95%CI | 1.032~1.408 | 0.871~1.226 | 0.941~1.531 | 0.625~0.870 |

CI, confidence interval; OR, odds ratio; \**P* value was calculated by 2-tailed $\chi^2$

## Notes

### Competing Interest Statement

The authors have declared no competing interest.

### Funding Statement

Not Applicable

